# “Getting pregnant during the COVID 19 was a big risk because getting the help from the clinic was not easy”: COVID-19 experiences of women and healthcare providers in Harare, Zimbabwe

**DOI:** 10.1101/2023.08.11.23293472

**Authors:** Zivai Mupambireyi, Frances Cowan, Elizabeth Chappell, Anesu Chimwaza, Ngoni Manika, Catherine J Wedderburn, Hannah Gannon, Tom Gibb, Michelle Heys, Felicity Fitzgerald, Simbarashe Chimhuya, Diana Gibb, Deborah Ford, Angela Mushavi, Mutsa Bwakura-Dangarembizi

## Abstract

**Background:** The COVID-19 pandemic and associated measures may have disrupted delivery of maternal and neonatal healthcare services and reversed the progress made towards dual elimination of mother-to-child transmission of HIV and syphilis in Zimbabwe. This qualitative study explores the impact of the pandemic on the provision and uptake of prevention of mother-to-child transmission (PMTCT) services from the perspectives of women and maternal healthcare providers.

**Methods:** Longitudinal in-depth interviews were conducted with 20 pregnant and breastfeeding women aged 20-39 years living with HIV and 20 healthcare workers in two maternity polyclinics in low-income suburbs of Harare, Zimbabwe. Semi-structured interviews were held after the second and third waves of COVID-19 in March and November 2021 respectively. Data were analysed using the modified grounded theory approach.

**Results:** While eight antenatal care contacts are recommended by Zimbabwe’s Ministry of Health and Child Care, women reported only being able to access two contacts. At antenatal booking, women were told to return at onset of labour; subsequent visits were suspended. Healthcare workers reported this reduction in antenatal attendance was a result of limited availability of personal protective equipment and fear that patients and services providers would contract SARS-CoV-2. Although HIV testing, antiretroviral therapy (ART) refills and syphilis screening services were accessible at first contact, other services such as HIV-viral load monitoring and enhanced adherence counselling were not available for those on ART. Closure of clinics and shortened operating hours during the second COVID-19 wave resulted in more antenatal bookings occurring later during pregnancy and more home deliveries. Six of the 20 interviewed women reported giving birth at home assisted by untrained traditional midwives as clinics were closed. Babies delivered at home missed ART) prophylaxis and HIV testing at birth despite being HIV-exposed. Although women were faced with multiple challenges, they continued to attempt to access services after delivery.

**Conclusions:** The COVID-19 pandemic disrupted provision and uptake of PMTCT services; antenatal care contacts were significantly reduced, home deliveries increased, and babies born at home missed out on the necessary ARV prophylaxis. These findings underline the importance of investing in robust health systems that are able to respond to emergency situations to ensure continuity of essential HIV prevention, treatment, and care services.

**Summary box:** *What is already known on this topic:* Studies have shown that the COVID-19 pandemic and associated control measures have disrupted provision of maternal and neonatal healthcare services globally.

*What this study adds:* The COVID-19 pandemic disrupted provision and uptake of PMTCT services; antenatal care contacts were significantly reduced, home deliveries increased, and babies born at home missed out on the necessary ARV prophylaxis.

*How this study might affect research, practice, or policy:* Our findings underline the importance of investing in robust health systems that are able to respond to emergency situations to ensure continuity of essential HIV prevention, treatment, and care services.

## Introduction

Zimbabwe recorded its first SARS-CoV-2 on 20th March 2020 and by November, 30, 2022, Zimbabwe had recorded 257,893 cases and 5,606 deaths [1] which is likely a huge underestimation due to limited testing coverage. The government of Zimbabwe instituted a number of policy, institutional and operational measures to contain community transmission of SARS-CoV-2 and prepare the fragile health system for an upsurge of cases [2, 3]. Public movement was restricted, gatherings were prohibited, informal markets, schools and other services were closed except those considered essential [4]. While these measures sought to curb the spread of SARS-CoV-2, experience from West Africa suggest that healthcare systems are broadly adversely affected by pandemics [5, 6]. In Guinea and Sierra Leone, maternal and child health indicators worsened substantially during the 2014 Ebola outbreak, with recovery still incomplete years later [7].

At the onset of the COVID-19 pandemic in Zimbabwe a number of demand and supply challenges were reported which disrupted uptake of health services. Reassignment of medical and laboratory staff, reallocation of resources, closure of healthcare facilities and insufficient personal protective equipment (PPE) resulted in reduced availability of clinical care in some settings, compounded by lack of availability of public transport which prevented health care staff getting to work [8, 9]. Healthcare workers went on strike due to the lack of PPE further compromising patients’ access to care [10]. Decreased care seeking was reported due to difficulty in navigating police road-blocks coupled with fear, mistrust and financial difficulties during lockdowns [11].

Emerging evidence from low-income countries suggests severe COVID-19 related disruptions to healthcare in early 2020 [11, 12]. In 2020 the GlobalFund reported widespread disruptions to HIV, TB and malaria service delivery, impacting approximately three-quarters of HIV, TB and malaria programs [12]. A rapid assessment conducted in Zimbabwe between March and April 2020 found that about 19% of people with HIV were unable to get ART [13], and similar data have been reported in other African countries[14–17]. Disruptions in HIV treatment and care services threatens the remarkable progress made towards elimination of HIV transmission, morbidity, and mortality. As pandemics disrupt health systems and affect human health globally it becomes important to understand the impact the COVID 19 had on the provision and uptake of Prevention of Mother-to-child Transmission (PMTCT) of HIV and syphilis services.

Zimbabwe has registered notable declines in HIV prevalence in the last decade, but continues to have one of the highest HIV burdens in Southern Africa [18]. HIV prevalence is estimated at 11.8% among women and men aged 15-49 years [18] and 16.7% among pregnant women [19, 20]. In 2011, Zimbabwe scaled up an accelerated national PMTCT program to eliminate paediatric HIV. The PMTCT services are integrated within all the public health facilities (1560) providing antenatal care (ANC) and maternal and child health services and Zimbabwe is targeting dual elimination of HIV and syphilis by Dec 2025 [21, 22].

In 2013 the Ministry of Health and Child Care (MoHCC) adopted the WHO guideline which recommended the provision of lifelong Antiretroviral Therapy (ART) to all HIV positive pregnant and breastfeeding women (Option B+) [25, 26]. With the implementation of Option B +, pregnant women are encouraged to register for ANC services in the first 12 weeks of pregnancy and have at least eight antenatal contact visits [28]. Dual HIV and syphilis testing services are offered to all women through an “opt-out” strategy at the first ANC contact [29]. Women who test HIV positive and are already on ART at first ANC visit continue with their treatment and have HIV viral load monitoring every 3 months till delivery. ART naïve women are initiated on ART [27, 28, 30]. After delivery, the mother-infant pairs are followed-up at maternal child health clinics until 24 months or the cessation of breastfeeding [27]. HIV negative women are retested at 32 weeks gestation and 6 weeks post-delivery.

In 2018, an estimated 93% of all pregnant and breastfeeding women living with HIV received ART through the national PMTCT programme [22]. In 2019, HIV and syphilis antenatal testing coverage were 98% and 91% respectively. The risk of vertical transmission at 18 months postnatal was estimated at 15% in 2015 and had fallen to 7.8% in 2018 [22]. During 2019, 56% of HIV-exposed infants received early infant diagnosis (EID) [26].Although progress has been made in reducing vertical transmission of HIV and syphilis, there were some gaps in the provision of PMTCT services before the COVID-19 epidemic. The emergence of the COVID-19 pandemic is likely to have dramatically disrupted delivery of maternal and neonatal healthcare services across the country. We therefore conducted a qualitative study with pregnant and breastfeeding women, Health promoters (known as Village Health Workers in rural areas) and Healthcare workers (HCWs) to describe the impact of COVID-19 on provision and uptake of PMTCT services in urban low-income areas in Harare, Zimbabwe. Uptake of ANC in this study is defined as seeking ANC services from trained healthcare providers and remaining in care pre- and postnatally while provision of care is referring to the provision of a range of ANC services on time by trained healthcare providers. This qualitative study was part and parcel of wider mixed method study which retrospectively looked at the effects of COVID-19 key PMTCT indicators using programme (District Health Information System-2) and population level data (ZIMSTAT) [23] and analysis of the outcomes of HIV-exposed neonates using NeoTree data [24].

## Methods

### Study design, sampling, and recruitment

Longitudinal semi-structured in-depth face to face interviews were held after the peak of the second and third waves of COVID-19 in March and November 2021 respectively. The first interviews were conducted after the introduction of the COVID-19 stringent measures to capture the women, health promoters and healthcare provider’s perception of COVID-19. Repeat interviews with the same participants were conducted when COVID-19 measures were slightly eased.

The study was conducted in two City of Harare polyclinics which provide maternity services. Mabvuku and Kuwadzana polyclinics are located in low-income, densely populated, urban suburbs in Harare. The clinics were selected because they have some of the highest number of deliveries among the municipality-owned clinics in Harare [31].

Women living with HIV who were pregnant or had given birth in the preceding 12 months were purposively recruited. A list of all pregnant and breastfeeding women who were in care or had delivered between March 2020 and February 2021 was drawn from the ANC and delivery registers in the two clinics. From this a stratified purposive sample of 20 women was selected to ensure a wide range of experiences considering age, history and duration of ART use, parity, and outcome of delivery. Additionally, 10 health promoters and 10 HCWs were purposively recruited based on the years in service and age. They were followed up at the same timepoints with the pregnant and breastfeeding women. In Zimbabwe community-based health promoters act as liaisons between the health facility and the suburbs and are knowledgeable of various health issues within their communities, including the number of pregnant and breastfeeding women, children (alive and deceased) and women who have discontinued HIV care and treatment programmes. Healthcare workers employed by either of the two study clinics who provide direct healthcare services to pregnant women seeking ANC services were recruited. HCWs were included because of their integral role in the provision of PMTCT services and their insight into structural influences such as the availability and accessibility of these services at the health-facility level.

### Data collection

All the participants had two repeat interviews conducted eight months apart. Data were collected by the first author and a trained research assistant using a semi-structured interview guide. All the interviews were conducted in Shona, the most widely spoken local language in Harare. Interviews lasted between 40 to 60 minutes. Interviews adhered to the MoHCC-guidelines on prevention and control of COVID-19 which included handwashing, wearing of masks, social distancing, and use of well-ventilated private rooms.

The first interviews conducted at the peak of the second COVID-19 wave in March 2021 explored retrospective experiences of being pregnant and giving birth during the first COVID-19 wave when there was little knowledge and severe preventative measures had been introduced. The interviews also explored current experiences during the second wave when restrictions were being relaxed. The second interviews were conducted in November 2021 before the subsequent surge in COVID-19 driven by the omicron variant when COVID-19 restrictions were relaxed, and information had become more widely available.

### Data analysis

All audio-recorded data were transcribed and translated verbatim into English. A coding framework was developed by the first author and an experienced research coordinator. Transcripts were then uploaded, coded, and summarized using a qualitative software package (NVivo 9.0, QSR International). Data were analysed using the modified grounded theory approach. In the initial stages we explored emerging themes and then adopted a constant comparison approach between different participants and clinics. The findings are presented under two main themes: uptake and provision of maternal and child health services. The perspectives of the women, health promoters and HCWs are presented together.

### Ethical Considerations

This study was granted ethical approval by the Joint Parirenyatwa Hospital and College of Health Sciences Research Ethics Committee (JREC 196/2020) and the Medical Research Council of Zimbabwe (MRCZ/A/2682). Approval to conduct the study was also granted by the MoHCC and City of Harare Health Department. All participants provided written informed consent and participation was voluntary.

### Patient and Public Involvement

Patients and/or the public were not involved in the design, or conduct, or reporting of this research. Patients and the public were however involved in the dissemination plans of this research as they outlined their preferred dissemination platforms and format.

## Findings

### Socio demographic characteristics of women, health promoters and healthcare providers

Twenty women aged between 20-39 years were recruited. Of these, 17 were married and staying with their partners, while two were not married and one was widowed during the course of her pregnancy. The widow’s partner had refused to be tested for HIV prior to the COVID-19 pandemic and eventually died of HIV-related complications. Twelve were informal traders, seven were unemployed and only one was formally employed. Sixteen women were breastfeeding while four were still pregnant at first interview. Among the breastfeeding mothers, their infants were aged between one to nine months while the pregnant mothers ranged between 13- and 28-weeks gestation. Nineteen had been infected with HIV later in life while one was perinatally infected. Seventeen women knew their HIV status prior to pregnancy while three were diagnosed following HIV testing at the first ANC contact and were immediately initiated on ART. Of the seventeen who knew their HIV status, fifteen were initiated on ART prior to pregnancy and two were not on ART despite knowing their HIV status. Denial of HIV status and fear of commencing ART were cited as the reasons for refusing ART. Of the seventeen that were married, fifteen knew their partner’s HIV status and two did not. Of the twenty women, six were first time mothers while fourteen already had children ranging from two to five children.

Health promoters were aged between 30-64 years, four had completed secondary level education, two had partial secondary education while four had primary level education. All the health promoters had been working in the communities for more than five years. Among the 10 healthcare workers five were midwives and five were registered general nurses.

### Uptake of maternal and child health services

Interviewed women, health promoters and HCWs talked about COVID-19 in terms of the challenges they experienced in accessing or providing healthcare services at family, community, and health facility level.

#### Fear of COVID 19 infection

At the family level, fear of contracting SARS-CoV-2 infection emerged as one barrier that deterred women from seeking ANC services. Women mentioned that they feared the highly infectious nature of the virus and that it was “uncurable and killing people in their thousands”. They feared failing to get the best possible care to help manage the infection if they got infected. Women anticipated devastating effects at family and community levels judging from the high mortality rates they were seeing on various news channels from high income countries. They had to take precautionary measures which included minimizing their movements and interaction with people. Their fears were compounded by the myths and misconceptions circulated on social media platforms at a time when people had limited information about the pandemic.

> “Fear gripped me, after I heard the number of people who were dying in countries abroad such as Italy. I have never heard of people dying in substantial numbers like that before and I imagined what it was going to do in a poor country like Zimbabwe and my poor community and I got so scared to such an extent that I did not want anyone near me or my family members. I was afraid to go to the shops, or to the borehole or even the clinic, I feared going out of my house as I could see COVID-19 all over in the air in people’s hands. I did not want anyone coughing near me I can say I was so scared” (Participant 1, woman, in her 30s).

> “I was afraid of COVID-19 because I heard that it was killing people with underlying health conditions like HIV. People were showing us videos of how people were getting infected in hospitals, and I imagined being in the same waiting area with someone with COVID or being attended to by a nurse with COVID and I was so scared and I told myself I will only go if there is a problem” (Participant 2 woman in her 20s).

During the first interviews which were conducted in March 2021, HCWs reported a decline in the number of women seeking ANC services due to a number of factors including fear of SARS-CoV-2 infection. Community members heard that HCWs had contracted SARS-CoV-2 and were afraid to come to the facility as one HCW mentioned.

> “We have seen a decline in the number of patients, people are not coming as they used to do because staff tested positive and the community heard about it, and so maybe they are fearing that maybe when they come here, they might eh contract COVID-19” (HCW 1).

Health promoters also spoke about the negative reception they received during home visits leading them to suspend the community follow ups. This is what they said:

> “We stopped doing home visits before the government announced the lockdown because people did not want us to come into their homes. We would come across notices at the gates or doors saying, ‘do not enter we do not want visitors here’. Many people in the community are still afraid of the virus considering how it is spread so they are not entertaining any visitors (Health promoter 1).

This fear was not limited to the women; health promoters and HCWs acknowledged being afraid because of their increased exposure to and risk of contracting of SARS-CoV-2. Looking after family members with underlying health conditions increased their fears.

> “It affected me a lot because I am a known diabetic and hypertensive patient, and I was afraid and anxious that if I go to work what if I get infected with the disease… some of my colleagues got infected and tested positive though with minor symptoms so I was worried about doing home visits that maybe I will get the infection from there, so it was difficult for me to carry out my usual duties” (Health promoter 2).

> “When the COVID started I was very afraid of getting the infection. I worried about getting the infection and passing it on to my mum who is asthmatic. I would dread coming to work and my fear worsened when my colleagues tested positive. As part of the mitigation measures, we resorted to use open spaces for our consultation since our rooms are very small and have poor ventilation” (HCW 2).

During the follow up interviews in November 2021, healthcare workers reported less fear of infection and that women were slowly re-engaging with health services, though the numbers had not yet reached pre COVID 19 levels.

> “We are now beginning to see them [women] coming though the numbers are not as before COVID-19. I guess it was the same with us [healthcare workers] we used to be afraid, but we are now used to working with clients and slowly getting back to our normal routine but at the same time being cautious since we don’t have adequate PPE, and we had 12 of our staff members who got infected” (HCW 3).

#### Fear of inadvertent HIV status disclosure to partners and other people

In addition to the fear of contracting COVID-19 women also feared inadvertent disclosure of their HIV status. Of the 20 interviewed women, six had not disclosed their HIV status to their partners. They faced challenges in collecting medication for themselves and their infants. The continued presence of their partners at home due to the travel restrictions made it difficult for these women to meet their scheduled ART refills and even to take their medication or administer the nevirapine prophylaxis to their babies. Prior to the COVID-19 pandemic some women would hide their medication and take it in the absence of their partners. One HCW stated that;

> “The issue of disclosure is a big issue we have so many cases of women who refused to disclose their status to their partners. So being in the same environment all the time, it was quite difficult, some were not taking well while others could not observe time but would randomly take at any opportunity which disrupted their routines. It was difficult and I think we will see the effects of COVID-19 on adherence soon enough” (HCW 4).

The COVID-19 pandemic also brought about confidentiality challenges for women accessing PMTCT services. Prior to the COVID-19 pandemic, clients coming for routine HIV treatment and care services had a designated space which afforded them more privacy. However, with the advent of the COVID-19 mitigation and prevention measures, patients had to queue outside the clinics and have their ART cards collected at the gate in the full view of other patients which inadvertently exposed their HIV status. During the first interview some women reported having stopped collecting their medication for fear of being stigmatised in their communities.

> “The difference now is that, if you come here, like some of us who are on ART, if you come you are told to wait at the gate, so we do not want to be seen by many people, but the way they are doing it nowadays makes it so difficult for us, everyone on the queue get to know about our statuses… We stand in the same queue but when they come to collect cards, they say that those who are on ART please hand in your cards and then they will call us one by one. I once left without getting served as my sister-in-law was also in the queue…” (Participant 3 woman in her 30s).

Health promoters also shared the same concerns about the inadvertent disclosure happening to patients queuing outside the clinics.

> “The clinic admin staff will go to the gate and collect the patients’ cards, and after they look for their green cards and check them. They then go back to the gate and start to call their names out whilst everyone is listening, and they will be a lot of people there. It’s like forced disclosure because you have no option because everyone else who will be at the gate will get to know that so and so is on ART. In the communities you hear them saying they will not come until COVID-19 is over but who knows when this will go away, and you can imagine what will happen to those mothers and their babies” (Health promoter 3).

Fear of inadvertent disclosure also made it difficult for health promoters to follow up on women who disengaged with HIV services or missed ART refills during the COVID-19 pandemic. The HCWs stated that before COVID-19, health promoters and Young Mentor Mothers would assist by tracking women living with HIV in the communities through home visits, but when COVID-19 came some women did not want to be followed up as they were staying with partners, in-laws, or relatives whom they had not disclosed to.

> “The challenge with some of these women is that they stay with their in-laws and other relatives who are not aware of their HIV status so it becomes difficult for these women to freely access services unlike before the pandemic when they could access services without telling anyone at home that they were going to the clinic. Now that everyone knows that movement is restricted, they would question their whereabouts” (HCW 2).

By the time of the follow up interviews, travel restrictions had eased, and free movement had resumed and women were able to access services at the clinics without using their ART cards as travel passes. Women reported that they were now able to collect their own ARVs or their infant’s medication without their partners and in laws knowing where they are going to. Health promoters also reported that they had resumed home visits though some women were still not allowing visitors to their homes and others still had their partners at home. Health promoters had also started educating and reassuring communities about the safety and the need to access essential services such as PMTCT services.

> “We have started conducting home visits and we are now going round educating people to go to the clinic when they have any health challenges. We have been assuring them that it is safe to seek services and that the clinics are doing all they can to protect patients from getting COVID-19” (Health promoter 4).

#### ANC services registration, follow up and infant monitoring

COVID-19 and its related control measures were reported to have affected ANC registration from both the uptake and supply side. At the family level, the COVID-19 pandemic and associated measures disrupted their social and economic lives which had a spiral effect on women’s uptake and utilization of health services. Closure of informal economy businesses including marketplaces and vending sites disadvantaged women from their only sources of livelihoods. Nine women stated that their partners lost income during the total shutdown while 12 women who were informal traders also lost opportunities to sell their wares. This affected household income, food security and ability to pay rent, resulting in some families relocating to their rural homes. Loss of income also impacted on their capacity to raise the USD25 required to register for ANC services within the City of Harare polyclinics. Five out of the twenty mothers that were interviewed reported registering their pregnancy after 28-weeks’ gestation because of the need to pay a registration fee and the closure of clinics. Registering after 12 weeks is considered late as per the MoHCC guidelines.

### Provision of maternal and child health service

Closure of clinics or reduced operating times resulted in women reporting difficulty with ANC registration, pre and postnatal care and facility delivery. During the first interviews, women narrated how difficult it was for them to register their pregnancy at the clinic, registration being the gateway to pre and postnatal care. However, despite all the challenges they encountered, women persisted trying to access PMTCT services for themselves and their babies. They repeatedly visited clinics to check if antenatal care and growth monitoring services had resumed.

> “I could not register on four times because for one to be among the 10 women they were taking per day you were supposed to be here by 4 am … and one of the guards told us that some women were paying for overnight accommodation at one of the houses next to the clinic. I ended up paying $7 USD to have accommodation and that’s how I managed to join the queue on time and got served” (Participant 4 woman in her 20s).

> “Things started to get slow at this clinic because of COVID-19, imagine we had to wake up at three in the morning to join the queue and then they say today we are not attending to clients, or they say that they are taking 5, or 10 people. So, it slowed service delivery so much. Like today I was here by 8 am but this is lunch time, and I am not yet served and after lunch they will be saying we are closing” (Participants 5 woman, in her 20s).

Although ART provision or initiation, HIV testing and syphilis screening services were accessible at first contact, other services such as HIV-viral load monitoring, enhanced adherence counselling and other diagnostic services such as ultrasound scan imaging and laboratory services were limited. Women reported that HCWs stopped doing the regular ANC examinations and reduced consultation time in a bid to promote social distancing and reduce the exposure risk as they had limited PPE. None of the 20 interviewed women managed all 8 of the recommended ANC contact visits; six managed five contacts, three had four contacts, one had three contacts, eight women managed two contacts while two women had their first contact at the onset of labour. Of the nine women who had four or more ANC contact visits some contacts were made before the second COVID-19 wave when facilities had not started implementing stern mitigation measures. At the peak of the COVID-19 women were told at ANC registration to only come back at the onset of labour as other subsequent monitoring visits were suspended in a bid to decongest the clinics.

> “Yes, getting pregnant was a big risk because getting the help from the clinic was not easy. When I came to register the pregnancy, I was turned away and only managed to register on the third attempt. After registering I was told to come back when I am due or if there is a serious need. They stopped all the other monthly ANC visits and even if you showed up for general consultation the nurses would just tell you to go back home so we missed some of the services” (Participant 6, woman in her 30s).

#### Closure of clinics

During the national lockdown, a number of maternity polyclinics, were closed in a bid to limit the spread of SARS-CoV-2 and concentrate limited resources on the pandemic. HCWs reported that in their experience the closure of polyclinics without prior notification led to an increase in home deliveries. Clinics were closed as HCWs contracted or were exposed to COVID-19 and they had to isolate or quarantine at home. One HCW commented.

> “We had to close, everyone was in isolation and they [mothers] had to go to neighbouring clinics but some of them didn’t have money to go to other clinics so, they delivered at home. Those mothers, you will find them maybe accidentally and ask why they do not have baby cards or maternity records then they will tell you that ‘I gave birth at home’ and has not been initiated on ART and the baby is 8 weeks old” (HCW 5).

Women who had booked for a facility delivery ended up giving birth at home assisted by untrained traditional midwives as a last resort as clinics within their catchment areas were closed. Six women who had planned a facility delivery ended up delivering at home. Women stated that their home delivery experiences were painful; some mothers narrated how they delivered at the clinic gate having been refused entry by the security guards as the clinic was closed.

Health promoters cited several incidences where mothers would call on them to assist them deliver their babies. One health promoter had this to say.

> “Countless times people were calling us to assist them to deliver their babies, but we also don’t know how to do it, so a lot ended up delivering on their own or with the help of traditional midwives. I know of a woman who reached a point of delivering by herself. The baby had a twisted umbilical cord and the baby died. She did not have the money to call an ambulance or even to look for someone to help, some of the traditional midwives were charging USD50” (Health promoter 5).

Babies who were delivered at home were at increased risk of getting infected with HIV as they either delayed staring or did not receive the recommended nevirapine prophylaxis and missed the HIV testing at birth despite being HIV-exposed. Most of the women also reported not receiving viral load testing throughout their pregnancy. Healthcare workers also confirmed that babies that were delivered at home missed essential PMTCT services. They stated that they are noticing babies coming for immunization that missed the nevirapine prophylaxis at birth and HIV testing at birth and at 6 weeks.

> “We are seeing a number of babies that missed nevirapine dose, the first 72 hours, or babies who were never tested for HIV and some of the conditions they were giving birth in at home were dirty environments and you see babies with all sorts of infections” (HCW 6).

HCWs also noted that they are beginning to see babies testing HIV positive at 6 weeks which had been rare. They stated that this could have been a result of the disruptions in viral load testing of pregnant mother, late HIV testing during pregnancy and babies missing ART prophylaxis.

> “We are beginning to record babies with an HIV positive result at 6 weeks which was now very rare. Before COVID-19 we would go for close to two or three months without recording any positives [babies testing positive] but now we are getting one of two per month and we all suspect it was because mothers failed to access services on time” (HCW 7).

#### Increased workload

Healthcare workers in the Kuwadzana clinic which remained fully operational throughout the various lockdowns and industrial action cited being overwhelmed by the increased number of women coming for services. HCWs mentioned that they had to suspend other services so that they could prioritise deliveries as they could hardly cope with the increased numbers coming from surrounding clinics. This was made worse by severe staff shortages as a result of staff needing to self-isolate following COVID-19 exposure. Additionally, some HCW took industrial action to protest shortage of PPE. Those HCWs at work, had to work long and overlapping shifts.

> “Sometimes we could not attend to all pregnant mothers, let’s say they are 50 who want to be helped and there are just only two nurses at ANC department because some were on strike because of lack of PPE and some were sick and some in isolation so at most they were two or three nurses attending to deliveries, reviews and other services. The surrounding clinics were closed, and all the mothers were coming here so, the nurses did what they could do under the circumstances and prioritised those in labour, but it was the most draining time…” (HCW 8).

HCWs faced difficulties getting to work due to the shortages of public transport. During the national lockdowns there was limited public transport, and no services provided for those working in essential services. Informal public transport operators were banned from operating during lockdown. This delayed HCWs’ arrival at work and affected clinic opening and closing times, thus further reducing the number of women seen per day. Frequent and often impassable police roadblocks/checkpoints compounded these issues.

> “Transport was a nightmare because there was so much pressure at the ZUPCO buses. You wait at the bus stop for 3 hours waiting for ZUPCO and you will get to work at 9 am and even going back home, it was the same so because of the shortage of transport so we were now working till 2pm so that we will have time to look for transport and get home before curfew time” (HCW 5).

Despite inadequate PPE, severe staff shortages and the increased workloads, HCWs tried their best to provide services. Interviewed HCWs narrated how they devised ways to escape police roadblocks so that they could get to work on time.

> “The police roadblocks were too many and quite frustrating I remember one day I was not allowed to pass even after telling them that I was a midwife. Its fortunate that I was driving, and I had to find an alternative route to get to work. Others on public transport had challenges sometimes they would just call to say I was not allowed to pass by the police and one person will attend to all the mothers which was very stressful” (HCW 9).

By the time of follow up interviews, services were starting to normalise, for example, HCWs reported that ANC contacts at their clinics had gone back to the recommended number and enhanced adherence counselling and viral load monitoring for women living with HIV infection had resumed. HCWs commented that the establishment of clear COVID-19 guidelines and standard operating protocols (SOPs) for infection prevention saw improvements in services provision and in the availability of PPE and the development of safety precautions. These improvements allowed resumption of the recommended ANC services. HCWs stated that they had undergone a number of trainings in the interim which improved their preparedness and safety. However, services such as physical examinations and infant growth monitoring were still suspended.

## Discussion

This study assessed the consequences of COVID-19 pandemic and associated measures on the uptake and provision of PMTCT services in two clinics in Harare, Zimbabwe. The study confirmed that provision of essential PMTCT services was disrupted mostly during the peak of the first COVID-19 wave. The study highlighted the fear of COVID-19 infection and fear of inadvertent HIV status disclosure and loss of income limited uptake of PMTCT services during the COVID-19 pandemic. At the facility level closure of clinics, staff shortages in part due to disruption in availability of public transport, reduced clinic operating times and number of patients seen per day and ultimately the uptake of services.

Pregnant and breastfeeding mothers stopped using healthcare services for fear of contracting infection or putting their families at risk. They worried that their healthcare facilities lacked capacity to manage people who got sick with COVID-19 infection and therefore wanted to avoid infection at all cost. This mirrors findings from other studies in Zimbabwe, South Africa, Liberia, Malawi, and Sierra Leone which also reported significant declines in the uptake of healthcare services due to fear of infection [4, 25–27]. An audit at central hospitals in Zimbabwe conducted in August 2020 found reduced utilisation of maternity services and a trend towards an increase in maternal mortality [2, 4]. In South Africa and Zambia, and Uganda fear of exposure to the SARS-CoV-2 prevented women from seeking healthcare services [26]. As a result, maternal and child health indicators worsened substantially during the COVID-19 pandemic. Our findings suggested that the fear was worsened by the limited knowledge about the infection, highlighting the need to invest in provision of timely accurate information to reassure the public and importance of accessing essential lifesaving services such as PMTCT. As part of this project, we developed and disseminated educational materials for pregnant women, their families and HCWs to mitigate the impact of COVID-19 on PMTCT services and uptake.

The study found that women had reduced access to the eight recommended antenatal care contact visits with the majority only being able to access two contact visits. Women struggled to register their pregnancies on time as clinics were either closed or had shortened operating hours. Poor emergency preparedness saw maternity clinics across the country closing including referral hospitals [2, 4] thereby limiting women’s access to services. We found that depending on timing, some women failed to register their pregnancy within the recommended 12 weeks of gestation despite having the registration fees, with two women having their first contact already in labour. This is in line with our quantitative findings which showed a substantial drop in initial ANC bookings early in the pandemic, although this subsequently recovered to pre-pandemic levels [23]. Fewer women receiving repeat HIV testing in pregnancy, which may have been because they had fewer contact visits. Late pregnancy registration has been shown to delay ART initiation for women who test HIV positive and are ART naïve. Delayed ART initiation, poor viral load monitoring during pregnancy and limited pre and postanal follow up lead to high attrition in PMTCT programmes [28, 29]. Studies conducted in Ethiopia [30], Ghana [31], Malawi [32] and Zimbabwe [28] showed that where they was low retention in the first six months in PMTCT programmes there have been higher HIV transmissions.

Analysis of the PMTCT cascade demonstrates that HIV transmission rates are driven by mother–infant pairs who fall out of the cascade at later stages [33, 34]. In our study some women had to give birth at home due to clinic closures and their HIV-exposed babies missed out on ARV prophylaxis and HIV testing which again increased the risk of MTCT. The quantitative findings show a reduction in the number of infants who received HIV testing from 27% to 18% during COVID-19 [23]. The gaps healthcare workers described in the provision of PMTCT services due to COVID-19 appear to have increased the rate of MTCT as HCWs are beginning to pick babies who are seroconverting. In Zimbabwe the success of PMTCT programs has always leveraged on high ANC contact visits and facility-based deliveries which mean a reduction in contact visits and facility based is likely to increase the risk of transmission. The findings point to the need to invest in robust health-systems that respond to emergency situations and ensure continuity of services for women throughout pregnancy and breastfeeding [28].

Despite the challenges both women and HCWs were very resourceful. At least some women were able to navigate the difficulties in accessing services. The HCWs we spoke to did their best to provide services under difficult circumstances. Despite their fears of COVID-19 infection, colleagues testing positive, limited PPE and transport challenges HCWs came up with strategies to continue service provision. A number of studies have highlight HCWs being very resourceful in emergency situations. In West Africa HCWs worked hard to provide healthcare services during the Ebola outbreaks despite being at highest risk of contracting the Ebola virus [35, 36].

The findings highlight the need for creating public awareness on the importance of seeking health services before introducing stern preventive measures that has a potential to limit access to healthcare services. Introduction of national lockdowns must be preceded with awareness campaign through a number of platforms to educate both the literate and illiterate members of the public on how to access services and the precautionary measures to take. Further research is needed to understand whether health care services have recovered to pre-pandemic levels and, if not, in which areas in order to inform MoHCC on areas that need further support.

## Conclusion

The COVID-19 pandemic disrupted provision and uptake of PMTCT services in two clinics in Harare, Zimbabwe. Antenatal care contact visits were significantly reduced, some women were being forced to give birth at home and their babies subsequently did not receiving the necessary ART prophylaxis. These findings have shown the importance of investing in robust health-systems that respond to emergency situations while ensuring continuity of provision of essential HIV prevention, treatment, and care services.

## Competing interests

The authors declare no conflict of interest. The funders had no role in the design of the study; in the collection, analyses, or interpretation of data; in the writing of the manuscript, or in the decision to publish the results.

## Data Availability

The data underlying this analysis are available as a supplementary file.

## Acknowledgements

We acknowledge support we received from the healthcare workers in Mabvuku and Kuwadzana clinics during data collection. We are grateful to all the participants who willingly took part in this study.

## Funding

This project was funded by ViiV Healthcare. The MRC Clinical Trials Unit at UCL is supported by the Medical Research Council (programme number MC_UU_00004/03). Felicity Fitzgerald was supported by an NIHR Development and Skills Enhancement Award.

## Notes

### Competing Interest Statement

The authors have declared no competing interest.

### Author Declarations

This study was granted ethical approval by the Joint Parirenyatwa Hospital and College of Health Sciences Research Ethics Committee (JREC 196/2020) and the Medical Research Council of Zimbabwe (MRCZ/A/2682).

